# Development and validation of the TabCAT-EXAMINER: tablet-based executive functioning factor score for research and clinical trials

**DOI:** 10.1101/2024.10.23.24315997

**Authors:** Mark Sanderson-Cimino, Katherine L. Possin, Dan M. Mungas, Emily W. Paolillo, Breton M. Asken, Elena Tsoy, Sabrina Jarrott, Yann Cobigo, Rowan Saloner, Kaitlin B. Casaletto, Ciaran Considine, Julie A. Fields, Joie Molden, Katya Rascovsky, Sandra Weintraub, Bonnie Wong, Hilary W. Heuer, Leah K. Forsberg, Julio C. Rojas, Lawren VandeVrede, Peter Ljubenkov, Gil D. Rabinovici, Maria L. Gorno-Tempini, William W. Seeley, Bruce L. Miller, Bradley F. Boeve, Howard J. Rosen, Adam L. Boxer, Katherine P. Rankin, Joel H. Kramer, Adam M. Staffaroni

## Abstract

**Objective:** The National Institute of Health (NIH) Executive Abilities: Measures and Instruments for Neurobehavioral Evaluation and Research (EXAMINER) is a validated laptop-based battery of executive functioning tests. A modified tablet version of the EXAMINER was developed on the UCSF Tablet-based Cognitive Assessment Tool (TabCAT-EXAMINER). Here we describe the battery and investigate the reliability and validity of a composite score.

**Methods:** A diagnostically heterogeneous sample of 2135 individuals (mean age=65.58, SD=16.07), including controls and participants with a variety of neurodegenerative syndromes completed the TabCAT-EXAMINER. A composite score was developed using confirmatory factor analysis and item response theory. Validity was evaluated via linear regressions that tested associations with neuropsychological tests, demographics, clinical diagnosis, and disease severity. Replicability of cross-sectional results was tested in a separate sample of participants (n=342) recruited from a Frontotemporal dementia study. As this separate sample also collected longitudinal TabCAT-EXAMINER measures, we additionally assessed test-retest reliability and associations between baseline disease severity and changes in TabCAT-EXAMINER scores.

**Results:** The TabCAT-EXAMINER score was normally distributed, demonstrated high test-retest reliability, and was associated in the expected directions with independent tests of executive functioning, demographics, disease severity, and diagnosis. Greater baseline disease severity was associated with faster longitudinal TabCAT-EXAMINER decline.

**Conclusions:** The TabCAT-EXAMINER is a tablet-based executive functioning battery developed for observational research and clinical trials. Performance can be summarized as a single composite score, and results of this study support its reliability and validity in cognitive aging and neurodegenerative disease cohorts.

## Introduction

Executive functioning is an umbrella term for a multidimensional set of cognitive processes that influence goal-directed behavior (Karr et al., 2018; Rabinovici et al., 2015). It is supported by a dispersed but connected set of brain regions (Alvarez & Emory, 2006; Bettcher et al., 2016; Kramer et al., 2014; Niendam et al., 2012; Reineberg & Banich, 2016) and is impacted by neurological conditions including epilepsy (Longo et al., 2013), multiple sclerosis (Witt & Helmstaedter, 2015), traumatic brain injury (Broadway et al., 2019), and neurodegenerative diseases such as Alzheimer’s disease and frontotemporal dementia (FTD) (Rabinovici et al., 2015; Staffaroni, Bajorek, et al., 2020). The parcellation of executive functioning into subdomains varies by study, but cognitive neuroscience and statistical approaches commonly identify subdomains such as inhibition, set-shifting, fluency/generativity, and working memory (Kramer, 2014; Miyake et al., 2000). The assessment of executive functioning is complicated by the breadth of cognitive tests used to measure these domains, many of which focus on a specific subcomponent (e.g., inhibition) (Faria et al., 2015). As a result, clinics and research studies differ in what aspect of executive functioning they are describing, which may attenuate reproducibility and hamper cross-study comparisons. To address these challenges, the National Institute of Health (NIH) funded the Executive Abilities: Measures and Instruments for Neurobehavioral Evaluation and Research (EXAMINER) project to develop a laptop-based battery of executive functioning tests that was informed by cognitive neuroscience, and could enable measurement of executive functioning in clinical trials and observational research (Kramer, 2014; Kramer et al., 2014). The project aimed to create a modular, flexible battery that captured the subcomponents of executive functioning. NIH-EXAMINER tasks of working memory, verbal fluency, and cognitive control were modeled using confirmatory factor analysis and item response theory (IRT) to create an Executive Composite score that summarized executive functioning into a single pragmatic and psychometrically robust outcome (Bott et al., 2014; Possin et al., 2014).

Over the last decade, the NIH-EXAMINER and the Executive Composite score have been implemented and validated in a myriad of studies. For example, scores on the NIH-EXAMINER are strongly associated with real-world executive behavior and with volumes of frontal, parietal, and subcortical brain regions known to support executive functioning (Possin et al., 2014). The NIH-EXAMINER has also shown utility for detecting and monitoring symptoms of neurological conditions including traumatic brain injury (Broadway et al., 2019), attention deficit disorder (Schreiber et al., 2014), and pediatric sickle cell disease(Schatz et al., 2014). The NIH-EXAMINER is sensitive to cognitive changes across a range of neurodegenerative diseases(You et al., 2014), and worse performance has been associated with greater disease severity, suggesting it measures clinically meaningful symptoms (Possin et al., 2014; Staffaroni, Bajorek, et al., 2020). Moreover, the Executive Composite score is a sensitive marker for early disease manifestations, as demonstrated by its ability to detect cross-sectional and longitudinal changes in presymptomatic FTD mutation carriers (Staffaroni, Bajorek, et al., 2020). The NIH-EXAMINER has been used to measure treatment effects on cognitive performance in pharmacological treatment trials (Assogna et al., 2021; Leite et al., 2022; Miller et al., 2014; Yu et al., 2018), including those targeting AD and other causes of mild cognitive impairment and mild dementia (Hajjar et al., 2020; Vossel et al., 2021), as well as non-pharmacological trials of mindfulness, cognitive training, and transcranial direct current stimulation (Assogna et al., 2021; Clark et al., 2021; Leite et al., 2022; Yu et al., 2018).

Since the development of the laptop-based NIH-EXAMINER, tablets have become a frequently used alternative to paper-and-pencil testing or laptops for cognitive assessments. Tablets offer similar benefits as computer-based testing (e.g., accurate logging of reaction time, automated scoring), but offer additional advantages with respect to portability, affordability, and user friendliness (Canini et al., 2014; Vaportzis et al., 2017). One such tablet-based testing platform is the UCSF Tablet-based Cognitive Assessment Tool (TabCAT; https://tabcathealth.com/). A subset of the validated NIH-EXAMINER tasks was programmed into the TabCAT platform (TabCAT-EXAMINER).

Here we describe the development and validation of the TabCAT-EXAMINER score. The precision of a composite score across a range of clinical presentations is improved when that score is developed in a heterogenous testing cohort (Kramer et al., 2014). As such, model development and initial validation analyses were conducted in a large, diagnostically heterogenous cohort of individuals with AD and related disorders, including FTD. Given the high frequency of executive dysfunction in FTD and the reported usefulness of the original NIH-EXAMINER in monitoring FTD (Miller et al., 2014), we selected participants from the ARTFL-LEFFTDS Longitudinal Frontotemporal Lobar Degeneration (ALLFTD) consortium with longitudinal TabCAT-EXAMINER data as a validation cohort. In addition to replicating findings observed in the development cohort, we conducted analyses in this validation cohort to further understand longitudinal performances, including test-retest reliability. We hypothesized that a single, general TabCAT-EXAMINER score will 1) fit the data well; 2) demonstrate convergent and divergent validity with other independent cognitive tasks; 3) be sensitive to a variety of neurodegenerative syndromes and associated with worsening disease severity; and 4) be sensitive to subtle cognitive changes in presymptomatic carriers of autosomal dominant FTD mutations (Staffaroni et al., 2022).

## Materials and Methods

### Participants

All participants completed at least one of the five tablet-based TabCAT-EXAMINER measures. Some participants were previously exposed to the verbal fluency measures which are administered as part of the standard Uniform Data Set (UDS) Version 3.0 neuropsychological battery, but baseline data represents the first time that participants completed one of the TabCAT-EXAMINER measures. Inclusion criteria required English as the primary language to remove variance associated with language that may impact performance on these tasks. An associated study is underway to evaluate the TabCAT-EXAMINER in a Spanish speaking cohort (see future directions). Written informed consent was obtained for all participants or their legal representative. Studies were approved by the Institutional Review Boards Data at UC San Francisco and Johns Hopkins (University Trial Innovation Network) in accordance with university institutional guidelines and the Helsinki Declaration.

The total sample comprised 2135 participants (Table 1). A subset of these participants were used as the developmental sample (n=1793), which included individuals recruited through the University of California, San Francisco (n=509) and the ALLFTD research consortium (n=1284), a multi-center study of FTD (Boeve et al., 2020). Similar to the original NIH-EXAMINER development sample (Kramer et al., 2014; Possin et al., 2014), this development subsample was symptomatically heterogeneous and syndromic diagnoses were made in consensus conferences using published criteria (Barker et al., 2022; Brooks et al., 2000; Dubois et al., 2007; Gorno-Tempini et al., 2011; Jack Jr et al., 2018; Litvan et al., 1998; McKeith et al., 2017; Neary et al., 1998; Petersen et al., 2010; Petersen et al., 2013; Petersen et al., 2014; Postuma et al., 2015; Rascovsky et al., 2011; Sachdev et al., 2014). The sample included cognitively unimpaired individuals (CU; n=786), and patients with mild cognitive impairment (MCI; n=237), Alzheimer’s dementia (AD; n=115), behavioral variant FTD (bvFTD; n=167), corticobasal syndrome (CBS; n=29), logopenic variant primary progressive aphasia (lvPPA; n=28), nonfluent variant PPA (nvPPA (n=44), progressive supranuclear palsy (PSP; n=37), and semantic variant PPA (svPPA; n=49). There were 112 participants with non-neurodegenerative diagnoses (e.g., primary mood etiologies) and/or small subsamples (e.g., amyotrophic lateral sclerosis: n<10) labeled as “other”. The MCI group included those with mild cognitive deficits and/or mild behavioral changes irrespective of suspected etiology (Barker et al., 2022).

**Table 1:** Sample description and TabCAT-EXAMINER scores by CDR®+NACC-FTLD Global Score subgroups.

|  |  | CDR®+NACC-FTLD Global Score* |  |  |  |  |  |
| --- | --- | --- | --- | --- | --- | --- | --- |
|  | Full sample | 0 | 0.5 | 1 | 2 | 3 | Comparison |
| Demographics |  |  |  |  |  |  |  |
| Sample Size | 1793 | 656 | 326 | 302 | 130 | 10 | --- |
| Age | 64.22<br>(13.94) | 59.19<br>(16.60) | 66.87<br>(11.23) | 66.65<br>(8.92) | 66.49<br>(9.58) | 71.00<br>(8.76) | 0<0.5 |
| Education | 16.38<br>(2.77) | 16.84<br>(2.36) | 16.42<br>(2.78) | 15.86<br>(2.81) | 15.59<br>(2.91) | 16.38<br>(2.77) | 0>0.5; 1<2 |
| Male n (%) | 860<br>(48.4) | 274<br>(41.8) | 172<br>(52.8) | 183<br>(60.6) | 78<br>(60.0) | 860<br>(48.4) | 0<0.5 |
| Diagnosis n (%) |  |  |  |  |  |  |  |
| CU | 786<br>(49.0) | 573<br>(91.2) | 56<br>(18.1) | 12<br>( 4.0) | 6<br>( 4.7) | -- |  |
| MCI | 237<br>(14.8) | 20<br>( 3.2) | 147<br>(47.4) | 26<br>( 8.8) | 4<br>( 3.1) | -- |  |
| AD | 115<br>( 7.2) | -- | 11<br>( 3.5) | 72<br>(24.2) | 12<br>( 9.4) | -- |  |
| CBS | 29<br>( 1.8) | -- | 9<br>( 2.9) | 12<br>( 4.0) | 4<br>( 3.1) | 1 (10.0) |  |
| lvPPA | 28<br>( 1.7) | -- | 3<br>( 1.0) | 19<br>( 6.4) | 6<br>( 4.7) | -- |  |
| bvFTD | 167<br>(10.4) | -- | 25<br>( 8.1) | 65<br>(21.9) | 62<br>(48.4) | 9 (90.0) |  |
| nvPPA | 44<br>( 2.7) | -- | 12<br>( 3.9) | 23<br>( 7.7) | 6<br>( 4.7) | -- |  |
| PSP | 37<br>( 2.3) | -- | 5<br>( 1.6) | 18<br>( 6.1) | 10<br>( 7.8) | -- |  |
| svPPA | 49<br>( 3.1) | -- | 13<br>( 4.2) | 29<br>( 9.8) | 6<br>( 4.7) | -- |  |
| Other | 112<br>( 7.0) | 35<br>( 5.6) | 29<br>( 9.4) | 21<br>( 7.1) | 12<br>(9.4) | -- |  |
| TabCAT-EXAMINER |  |  |  |  |  |  |  |
| Factor Score | -0.03<br>(0.91) | 0.56<br>(0.61) | -0.19<br>(0.70) | -0.75<br>(0.73) | -1.06<br>(0.76) | -1.91<br>(0.62) | 0>0.5>1>2 |
| Flanker | 7.37<br>(1.54) | 8.09<br>(1.00) | 7.05<br>(1.41) | 6.35<br>(1.72) | 5.66<br>(1.55) | 5.46<br>(0.30) | 0>0.5>1 |
| Set Shifting | 7.04<br>(1.52) | 7.84<br>(1.02) | 6.56<br>(1.37) | 5.93<br>(1.70) | 5.29<br>(1.31) | 5.25<br>(1.21) | 0>0.5>1 |
| **Match** | 41.91<br>(13.80) | 50.71<br>(7.64) | 40.62<br>(10.41) | 31.12<br>(13.13) | 26.27<br>(14.04) | 22.70<br>(15.04) | 0>0.5>1>2 |
| **Dot Counting** | 15.23<br>(5.80) | 17.92<br>(4.54) | 14.51<br>(4.95) | 11.88<br>(5.41) | 9.80<br>(7.07) | 3.50<br>(3.15) | 0>0.5>1 |
| **Running Dots** | 29.65<br>(7.86) | 32.40<br>(6.77) | 26.38<br>(7.46) | 28.15<br>(8.11) | 25.82<br>(7.68) | 29.00<br>(NA) | 0.5>1 |
| **Animals** | 18.63<br>(8.97) | 24.90<br>(6.17) | 17.05<br>(7.01) | 12.31<br>(6.57) | 10.00<br>(5.87) | 3.75<br>(3.11) | 0>0.5>1 |
| **Vegetables** | 11.90<br>(6.25) | 16.04<br>(4.69) | 10.41<br>(4.57) | 7.90<br>(5.19) | 6.13<br>(4.31) | 2.88<br>(2.10) | 0>0.5>1 |
| **Words (F)** | 12.36<br>(6.43) | 16.26<br>(4.60) | 11.46<br>(5.41) | 8.25<br>(5.54) | 6.71<br>(4.88) | 2.38<br>(1.77) | 0>0.5>1 |
| **Words (L)** | 11.44<br>(5.78) | 14.69<br>(4.13) | 10.93<br>(4.90) | 8.02<br>(5.14) | 6.20<br>(5.08) | 2.75<br>(1.49) | 0>0.5>1 |

The "Comparison" column indicates significance of comparisons (T-test or Chi<sup>2</sup> test) between CDR®+NACC-FTLD Global score groups. Statistical comparisons involving the CDR®+NACC-FTLD Global Score = 3 group were not computed due to small sample size. Means, (standard deviations), and, when applicable, sample size [n], are presented. There were 368 participants missing CDR®+NACC-FTLD Global Scores.
CU: cognitively unimpaired; MCI: mild cognitive impairment; AD: Alzheimer's dementia; bvFTD: behavioral variant FTD; CBS: corticobasal syndrome; lvPPA: logopenic primary progressive aphasia; nvPPA: nonfluent variant PPA; PSP: progressive supranuclear palsy; svPPA: semantic variant PPA. Other: participants with non-neurodegenerative diagnoses (e.g., primary mood etiologies) and/or small subsamples (e.g., amyotrophic lateral sclerosis: n<10).

A different subset of ALLFTD participants (n=342) not included in the discovery cohort comprised the validation cohort. This subsample had longitudinal TabCAT-EXAMINER data, completed 6-24 months after their baseline (mean follow-up length: 368 days; SD=64). This sample included those diagnosed as: CU (n=140), MCI (n=21), AD (n=2), bvFTD (n=40), CBS (n=4), lvPPA (n=1), nvPPA (n=14), PSP (n=8), and svPPA (n=14). There were 13 participants who fell into the categories labeled as “other” in the developmental sample (e.g., primary etiology mood, n=3; Parkinson’s disease, n=1).

A description of the sample with genetic testing is presented in Supplemental Tables 1 and 2. DNA samples were screened using targeted sequencing of a custom panel of genes previously implicated in neurodegenerative diseases, including *GRN* and *MAPT*. Hexanucleotide repeat expansions in *C9orf72* were detected using both fluorescent and repeat-primed PCR (Ramos et al.). The developmental cohort included 79 participants who carried pathogenic mutations (*MAPT*, n=23; *C9orf72*, n=43; and *GRN*, n=13) known to cause familial frontotemporal lobar degeneration (f-FTLD). There were also 132 non-carriers from families with an identified pathogenic f-FTLD mutation (i.e., family controls). The validation cohort included 80 participants who carried f-FTLD genetic mutations (*MAPT*, n=24; *C9orf72*, n=44; and *GRN*, n=12). There were also 101 non-carriers from families with an identified pathogenic f-FTLD mutation in the validation cohort.

### TabCAT-EXAMINER

The TabCAT-EXAMINER comprises nine executive functioning tasks, including five tablet-based measures and four measures of verbal fluency. Tablet-based measures included tasks of inhibition (Flanker), set-shifting (Set Shifting), processing speed/working memory (Match), and working memory (Dot Counting, Running Dots) (Mullady et al., 2022; Possin et al., 2018; Weintraub et al., 2009). As noted below, the TabCAT-EXAMINER tasks were similar to the best performing measures of the original NIH-EXAMINER, with modifications to shorten tests, include more sensitive practice trials to reduce patient burden, and to add another brief, well-validated assessment of executive functioning (Match) already available through the TabCAT platform.

#### Flanker

The participant is presented with a display of arrows and asked to indicate which direction the central arrow is pointing. There are two conditions: in the congruent condition the non-target stimuli point in the same direction as the central arrow; in the non-congruent condition, the non-target stimuli point in the opposite direction of the central arrow. This task was modeled after the original NIH-EXAMINER Flanker task (Kramer et al., 2014). Minor alterations included the creation of practice trials; a reduction in total number of response trials; and slight changes in the minimum and maximum response times allowed. The primary outcome for this task is a score that combines speed and accuracy (range: 0-10); see (Mullady et al. 2022) and the original NIH-EXAMINER Flanker Task (Kramer et al., 2014; Weintraub et al., 2013) for details.

#### Set Shifting

Participants are asked to match a stimulus presented at the top of the screen to one of two stimuli at the bottom of the screen. In the task-homogenous condition, participants are asked to match on shape or color. In the task-heterogenous condition, participants are asked to alternate between matching color and shape, pseudo-randomly. Similar to Flanker, this task was modeled after the original NIH-EXAMINER Set Shifting task and underwent minor alterations for the TabCAT system. Similar to Flanker, the outcome variable combines accuracy and reaction time across all task-heterogenous conditions (range: 0-10) (Kramer et al., 2014).

#### Match

Participants are shown a set of numbers one to seven that are each matched to a simple shape in a key at the top of the screen. At the bottom of the screen the same set of shapes are presented. At each trial a number appears at the center of the screen. Participants are asked to tap the corresponding shape at the bottom of a screen as quickly as possible for two minutes. Like the classic Digit Symbol Substitution Test paradigm on which it was based, Match is a highly sensitive but not specific measure of executive functioning that relies on processing speed, attention, visual-motor coordination, and working memory (Jaeger, 2018).

#### Dot Counting

Participants are asked to count the total number of blue circles on a display. The number of circles ranges from two to nine per display, with additional distractors (e.g., squares). The number of displays presented increases each trial, and at the end of each trial, the participant is asked to recall the total number of blue circles per display. There are six total trials. This task is modeled after the original NIH-EXAMINER Dot Counting task and assesses verbal working memory (Kramer et al., 2014). The outcome variable is the total number of correct responses across all trials (range: 0-27) (Mullady et al., 2022).

#### Running Dots

Participants are shown a 16-square grid where a series of dots appear, one by one. They are asked to recall the last two to five locations where a dot appeared, after the dots disappear from the screen. The condition defining the target dots varies (e.g., the last two dots vs last four dots), as does the number of dots presented before the target dots. Participants are awarded a point for each dot they recall in the correct location and order; they are awarded half a point for recalling the correct location of a dot, but in the incorrect order. This task assessing spatial working memory was designed and implemented only in the TabCAT system. The primary outcome variable is the sum of total points across all trials (range: 0-42).

#### Verbal fluency

Measures included semantic fluency (Animals, Vegetables) and phonemic fluency (words [L], words [F]) collected as part of the UDS version 3.0 Neuropsychological Battery (Weintraub et al., 2018). Although these tasks are not administered on the tablet, the TabCAT system allows the user to input verbal fluency scores.

### Other assessments

#### Additional Cognitive Tasks

In addition to the TabCAT-EXAMINER, participants completed a battery of standard paper-and-pencil neuropsychological measures within 180 days of tablet testing (supplemental methods). Measures included four UDS executive functioning tasks (Lezak et al., 2004; Possin et al., 2011; Weintraub et al., 2009). Participants also completed tasks of visuospatial ability (Benson Figure copy), visual memory (Benson Figure Recall (Possin et al., 2011)), verbal memory (Craft Story Recall (Craft et al., 1996) and California Verbal Learning Test-II, Short Form (Delis et al., 2000)), and naming (Boston Naming Task (Weintraub et al., 2018) or Multilingual Naming Test (Stasenko et al., 2019)). Participants scores on the Multilingual Naming Task were converted to Boston Naming Task scores based on a prior crosswalk analysis (Monsell et al., 2016). Finally, participants completed two additional measures on the TabCAT platform that are not part of the EXAMINER: Line Orientation, a measure of visuospatial perception, and Favorites, a measure of associative memory (Possin et al., 2018).

#### Clinical Dementia Rating Scale Plus Behavior and Language domains from the NACC FTLD module (CDR®+NACC-FTLD)

The CDR®+NACC-FTLD (Knopman et al., 2008) is a clinician-administered, semi-structured interview with participants and informants that adds language and behavior domains to the original CDR (Morris, 1997) to account for core impairments of FTD-spectrum disorders. An algorithm (Miyagawa et al., 2020) was used to calculate a Global score that can be used to characterize disease severity as asymptomatic (0), prodromal (0.5), or symptomatic (1, 2, or 3). A sum of the eight domain Box Scores (CDR®+NACC-FTLD sum of boxes) was also calculated (max score = 24). Greater Global and Box Scores indicate more functional impairment (Miyagawa et al., 2019, 2020). This measure was administered to participants with and without FTD.

### Factor score modeling

Model building was conducted using the statistical software R (Team, 2021) and is fully described in the supplement. Briefly, exploratory factor analysis was used to determine the number of factors. A confirmatory model was then fit, followed by a multilevel IRT model using the MIRT package (Chalmers, 2012).

IRT is a form of latent variable modeling, similar to confirmatory factor analysis. This method differs from traditional factor analysis in that it models item difficulty as well as the item’s ability to discriminate between participants with different abilities. As a result, items in IRT models are weighted in how effectively they assess the target ability at varying levels along that ability continuum (Mungas et al., 2003). In the present analyses, we utilized a graded response model that allows for ordinal variables (Samejima, 2011) and recoded continuous variables into ordinal scores with up to 12 response categories and at least five observations in each category. This method retained the overall distribution of the raw, continuous data. The IRT model set the loadings within each fluency factor to be equal; this approach in MIRT is equivalent to specifying correlated measurement error in a CFA model. Acceptable model fit was determined via factor loadings (>.4 retained (Matsunaga, 2010; Williams et al., 2010)), the Tucker Lewis Index (TLI; >0.95 (Schumacker & Lomax, 2004)), the Comparative Fit Index (CFI; <0.90 (Savalei & Bentler, 2006)), and the Root Mean Square Error of Approximation (RMSEA; <.08) (Byrne, 2001).

A series of analyses were conducted to determine the reliability and utility of the TabCAT-EXAMINER score when calculated with fewer components, mimicking scenarios in which not all tests were administered. We retained the thresholds and loadings from the full model (described above) in a series of sub-models that retained two to seven components; all possible 502 combinations were evaluated. A separate factor score was generated by each model. Models were then grouped by number of components (e.g., two-component model, three-component model) and by requiring a specific component be included (e.g., two-factor models that included Match). To evaluate the reliability and internal consistency of the TabCAT-EXAMINER score, each reduced factor score was correlated with the full, nine-component TabCAT-EXAMINER. To evaluate the utility of a factor score generated with fewer than nine components, each reduced factor score was also correlated with the CDR+NACC FTLD sum of boxes. Results were separated by the number of components included in the model and by specific components.

### Convergent and divergent validity

To evaluate construct validity, we tested the association of the TabCAT-EXAMINER score with independent neuropsychological measures and scales of disease severity. First, a linear regression examined the association between the CDR®+NACC-FTLD Sum of Boxes and the TabCAT-EXAMINER score. Next, separate linear regressions were fit to assess the association between the TabCAT-EXAMINER score and age, education, sex, and neuropsychological measures while controlling for the CDR®+NACC-FTLD Sum of Boxes. Correlations did not adjust for demographic variables, as the goal was to understand the relationship between the TabCAT-EXAMINER score and paper-and-pencil measures rather than to evaluate the scores incremental utility above and beyond demographics. In a second set of analyses, linear regression was used to evaluate differences in TabCAT-EXAMINER scores across levels of clinical severity (CDR®+NACC-FTLD Global score), controlling for age, education, and sex. As we would expect executive functioning to differ between syndromic diagnostic groups and controls, we then demonstrated known group validity by assessing differences in TabCAT-EXAMINER scores across diagnostic syndromes through linear regressions, controlling for age, sex, and education.

Based on prior studies (Possin et al., 2014; Staffaroni et al., 2021), we hypothesized a similar degree of executive impairment across diagnostic groups. We next evaluated the ability of the TabCAT-EXAMINER to differentiate asymptomatic and prodromal f-FTLD mutation carriers (CDR®+NACC-FTLD Global score < 1; n=33) from family controls (CDR®+NACC-FTLD Global score = 0; n=56) using regression. Covariates included education, age, and sex. This analysis was restricted to those over the age of 45 to remove participants likely to be many years from symptom onset based on natural history studies (Staffaroni et al., 2022).

### Validation cohort and longitudinal analyses

#### Cross-sectional Analyses

Mirroring analyses in the development sample, regressions assessed the association between the TabCAT-EXAMINER score and age, education, sex, CDR®+NACC-FTLD Sum of Boxes, and neuropsychological measures. Regressions next compared TabCAT-EXAMINER scores across diagnostic syndromes. We then replicated comparisons between asymptomatic and prodromal f-FTLD mutation carriers (n=37) from family controls (n=31) using regressions in a sample over the age of 45.

#### Longitudinal Analyses

First, in a sample defined as functionally intact (CDR®+NACC-FTLD Global Score=0) at baseline and follow-up, test-retest reliability was assessed as the correlation between baseline and longitudinal scores. for Second, to test the hypothesis that rates of decline in TabCAT-EXAMINER scores hasten as disease severity increases, we fitted linear regressions with TabCAT-EXAMINER annualized change scores ([visit 2 – visit 1]/ retest-interval) as the outcome and baseline disease severity (CDR®+NACC-FTLD Global score group) as the predictor. Third, we fitted linear regressions to test whether those who converted to a higher CDR®+NACC-FTLD Global score at follow up showed more rapid TabCAT-EXAMINER declines than those who remained stable at follow up. Covariates included education, sex, and retest interval.

## Results

### Model building

The exploratory factor analysis scree plot (Supplemental Figure 1) suggested a one factor model accounted for the majority of variance (84%): eigenvalue_1-factor_: 5.53; eigenvalue_2-factor_: 1.0; eigenvalue_>2-factors_: mean= .35 (SD=.23). The CFA model, which accounted for shared measurement variance of the verbal fluency tasks, fit the data well (CFI=0.996, TLI=0.995, RMSEA=0.028) with moderate-to-strong factor loadings. This final model (Figure 1) was specified as a multilevel IRT model to generate factor scores.

**Figure 1:**
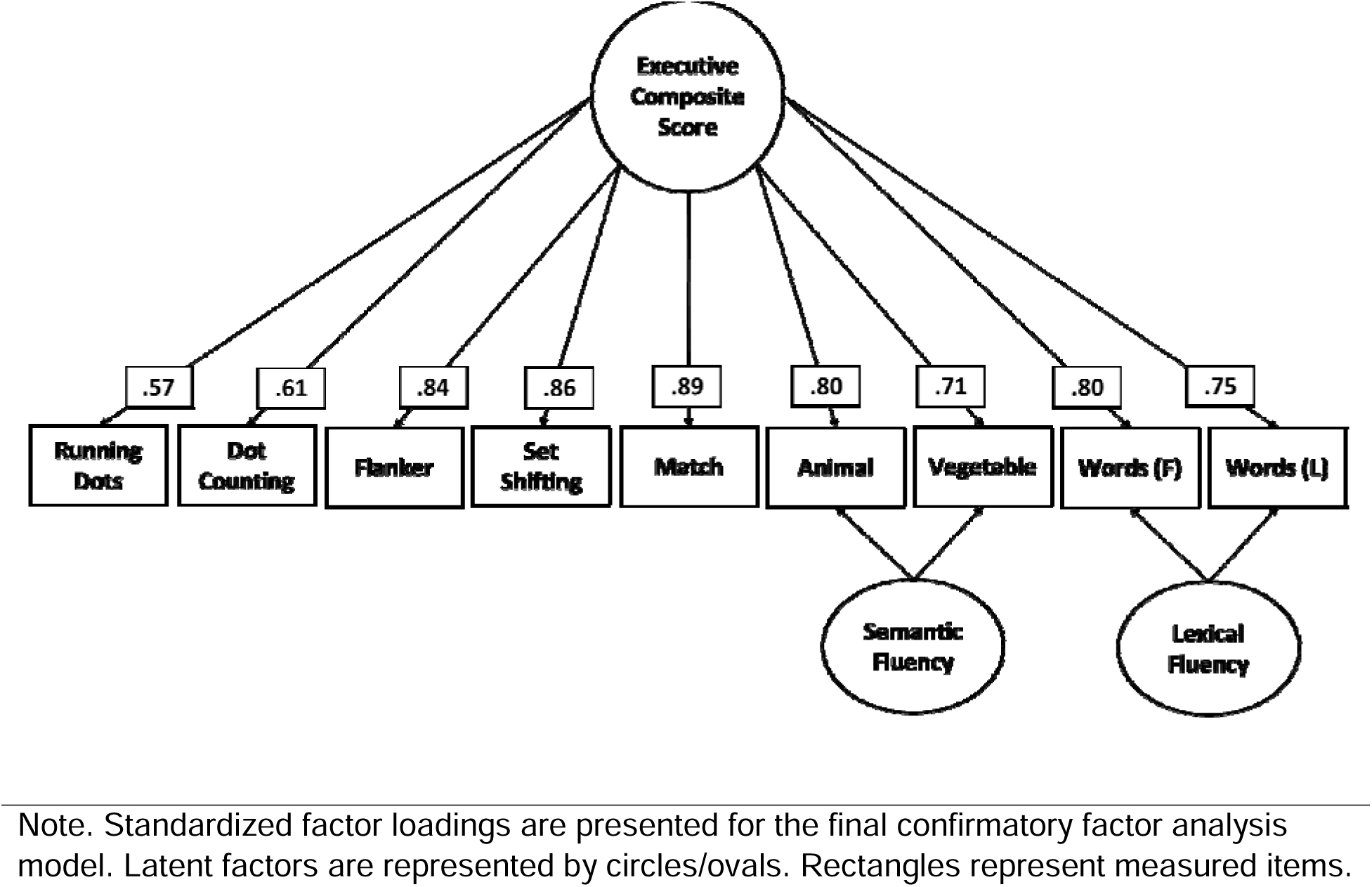
TabCAT-EXAMINER confirmatory factor analysis.

Factor score distribution and reliability are detailed in the supplement (Supplemental Figures 2-4). Briefly, the TabCAT-EXAMINER score was normally distributed. A series of models calculated the TabCAT-EXAMINER with fewer than nine components. As the number of components included in a model decreased, there was a reduction in the average correlation and an increase in variability around that association. However, the average correlation with the full TabCAT-EXAMINER remained strong, ranging from .84 to .90 across models. These results are presented in Supplemental Figure 3. The association of the reduced models and the full TabCAT-EXAMINER varied somewhat by specific component (Supplemental Figure 4), particularly when fewer than four components were included. There was also variability in the association between the reduced factor score and the CDR+NACC FTLD sum of boxes when fewer components were included (Supplemental Figure 5).

### Developmental sample: convergent and divergent validity

Associations between the TabCAT-EXAMINER score and paper-and-pencil neuropsychological tasks in the developmental sample are presented in Figure 2 (circles). Higher TabCAT-EXAMINER scores (better performances) were associated with more years of education (β =.09, [95%CI: .05, .13], p<.001) and younger age (β =-.35, [95%CI: –.38, –0.31], p<.001). Additionally, paper-and-pencil executive functioning tasks were significantly associated with the TabCAT-EXAMINER score in the expected direction, and standardized betas suggest large effects (β tasks of non-executive domains (i.e., language, memory, visuospatial) were also significantly associated with the TabCAT-EXAMINER score, with modest reductions in magnitude (β compared to the magnitude of associations with tasks of executive functioning.

**Figure 2:**
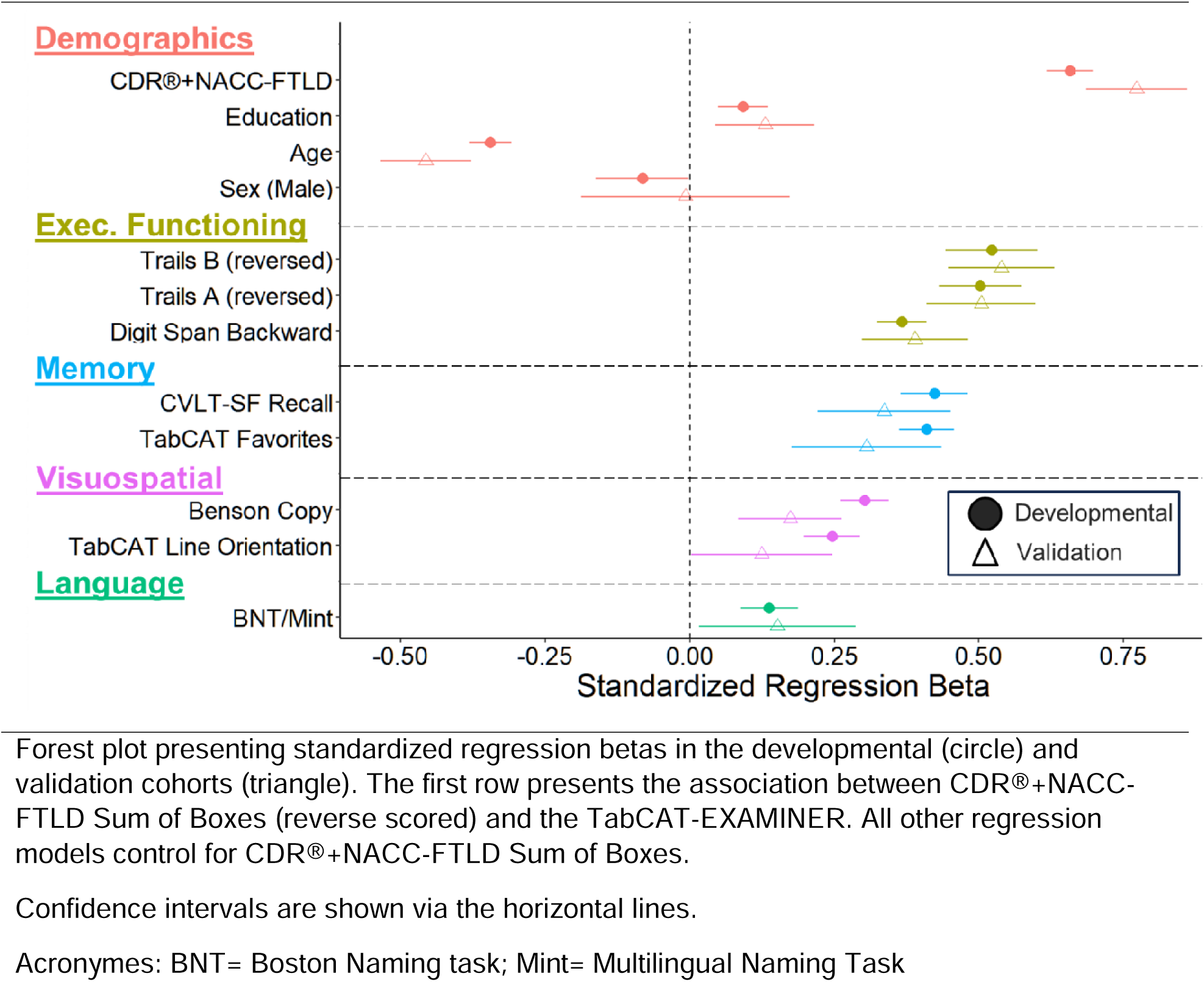
Associations of the TabCAT-EXAMINER score with demographics and clinical measures.

Lower TabCAT-EXAMINER scores were strongly correlated with greater disease severity as measured by the CDR®+NACC-FTLD Box Score (β =-.65, [95%CI: –.67, –.61], p<.001), and remained β =-.16, [95%CI: –.16, –.14], p<.001). There was a stepwise decline in factor score was noted with increasing CDR®+NACC-FTLD Global score (0 > 0.5 > 1 > 2; p<0.001). Table 1 provides summary scores by CDR®+NACC-FTLD global scores across all TabCAT-EXAMINER components and factor score.

The functionally intact control group had a significantly greater TabCAT-EXAMINER score than any of the symptomatic groups (Figure 3; *p*’s<.006). As a follow-up analysis, the control group was compared to symptomatic groups with at least 1 positive AD-biomarker. AD-biomarker positivity was based on PET-imaging, CSF, and/or plasma and diagnoses was determined in a consensus conferences using published criteria (Jack Jr et al., 2018). Results supported a stepwise decline in factor scores (*p*’s<.001) with increasing severity: cognitively unimpaired (n=647; mean=.53, SD=.63), mild cognitive impairment likely due to Alzheimer’s disease (1+ AD biomarker; n=40; mean=-.4; SD=.54), and Alzheimer’s disease dementia (1+ AD biomarker; n=37; mean=-1.13, SD=.74) groups.

**Figure 3:**
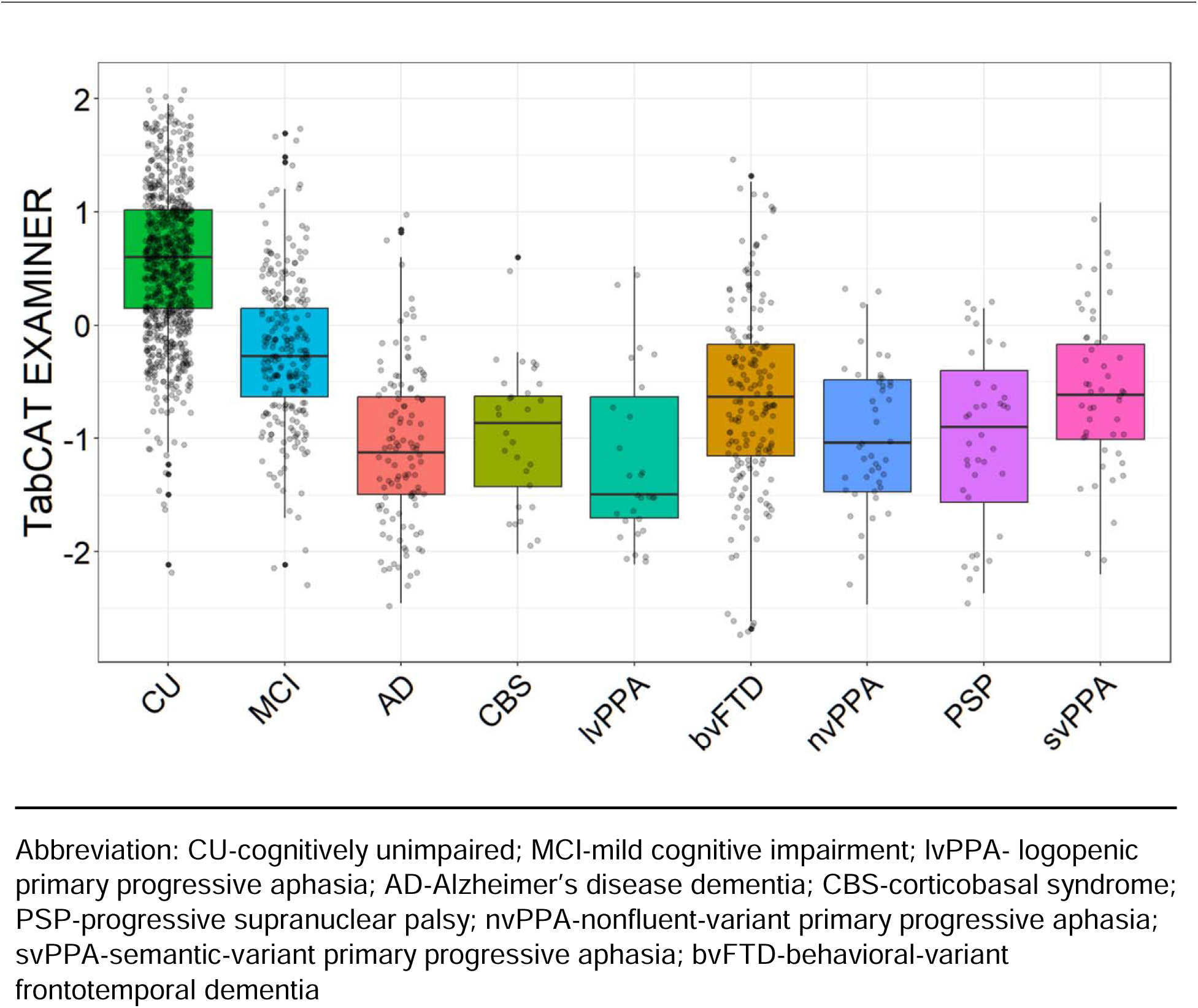
TabCAT-EXAMINER performance by clinical syndrome within the developmental sample.

In a subsample of individuals over the age of 45 (n=89), the average score for asymptomatic/prodromal f-FTLD mutation carriers was worse than family controls, although this difference was not statistically significant (β =-.21, [95%CI: –.43, –.01], p=.06).

### Validation sample and longitudinal analyses

Figure 2 provides associations among the TabCAT-EXAMINER score, demographics, and paper-and-pencil neuropsychological tasks in the ALLFTD validation sample (triangles). This figure demonstrates the similarities between standardized regression coefficients in the validation and development sample (e.g., association with Trail Making Test B: ALLFTD Validation: β = .54; Development: β = .52). EXAMINER score was correlated with baseline CDR®+NACC-FTLD® Box Score (β =-.74, [95%CI: –.78, –.70], p<.001),, and this association remained significant while controlling for age, education, and sex, (β =-.16, [95%CI: –.18, –.14], p<.001). Similar to findings in the developmental sample, a stepwise decline in factor score was observed with increasing CDR®+NACC-FTLD® Global score (0 > 0.5 > 1 > 2; p’s<.01). As found in developmental cohort, the majority of diagnostic groups performed significantly worse on the TabCAT-EXAMINER score than the cognitively unimpaired sample (i.e., CU > CBS, bvFTD, PPA, PSP, svPPA). Details are presented in Supplemental Figure 6. Within a small sample of asymptomatic/prodromal individuals from families known to carry f-FTLD mutations (n=68), those with mutations showed a trend towards worse performance on average than noncarrier family members on the baseline TabCAT-EXAMINER (β =-.17; 95%CI: –.38, .05; p=.13), although this difference was not statistically significant. Of note, this association was similar in magnitude to that observed in the development sample (β =-.17 vs –.21).

In those who remained unimpaired at follow up (n=146, mean interval=383 days (SD=52), test-retest reliability of the factor score was 0.86 (95% CI: 0.81, 0.90). Supplemental figure 7 provides information on test-retest reliability for all TabCAT-EXAMINER subtests. Annualized change scores were calculated for all participants. TabCAT-EXAMINER scores tended to decline more rapidly with more advanced disease progression (Figure 4). Examining the between-group differences, the asymptomatic group demonstrated significantly less annualized change than the prodromal group (β=-.16; 95% CI: –0.29, –0.03; p=.02) and the symptomatic group (β =-.22; 95% CI: –0.32, –0.15; p<.001). The comparison between prodromal and symptomatic groups was in the expected direction but the effect was small and not statistically significant (β =-.11; 95% CI: –0.29, –0.07; p=.23).

**Figure 4:**
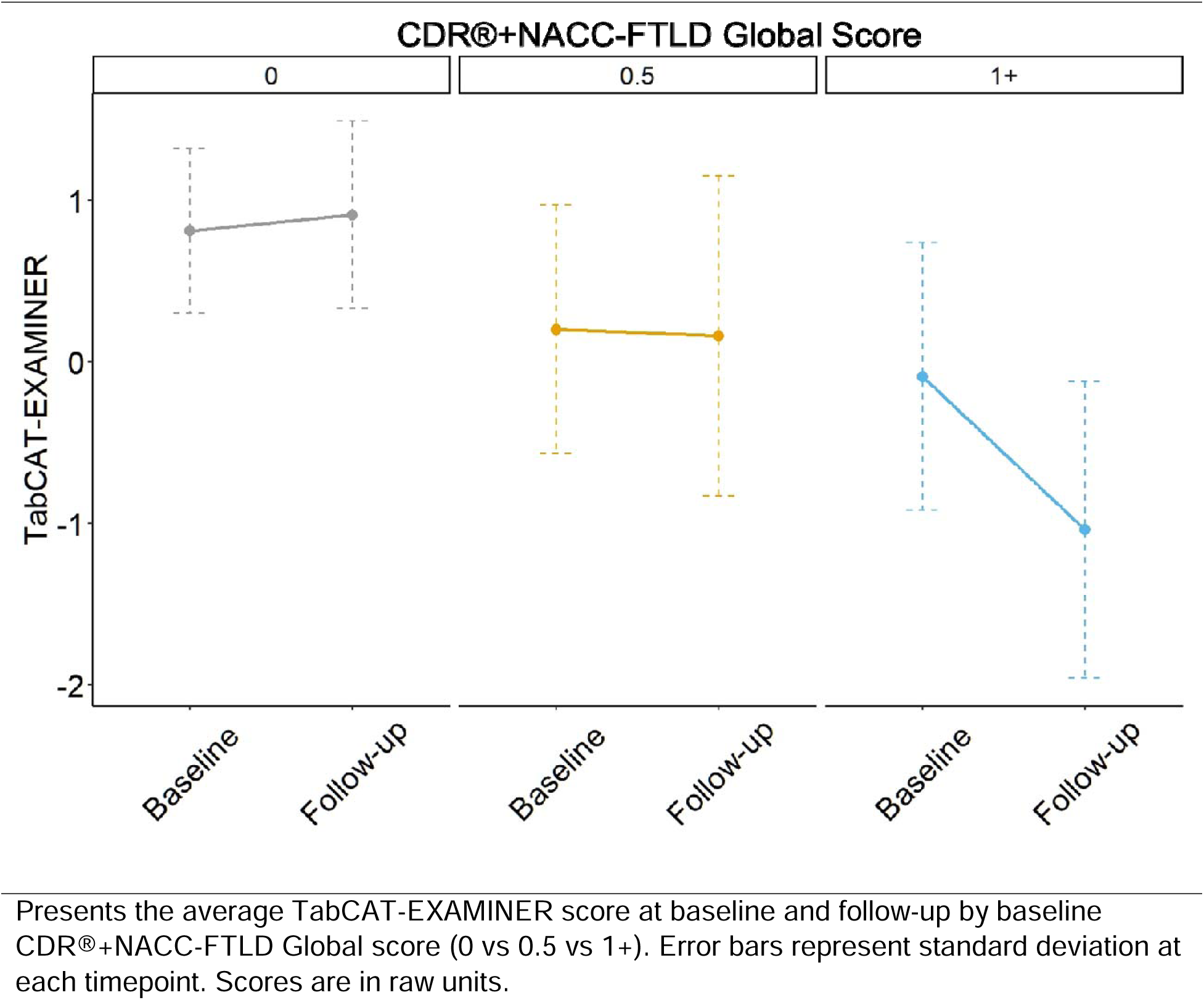
Longitudinal changes in TabCAT-EXAMINER scores by baseline disease severity.

Asymptomatic participants (CDR®+NACC-FTLD = 0) who progressed to show at least mild symptoms (CDR®+NACC-FTLD ≥ 0.5) at follow-up (n=14) showed significantly greater annualized decline (moderate effect size) in TabCAT-EXAMINER scores (β =-0.34; 95% CI: –0.52, –0.16; p<.001) than those who were unimpaired at both time points.

## Discussion

We present the development and validation of the TabCAT-EXAMINER score. Fit statistics indicated the model fit the data well, with moderate-to-strong factor loadings for all subtests, and the normally distributed factor score showed high test-retest reliability. Our data support the construct validity and psychometrics of the TabCAT-EXAMINER score through strong associations with other tasks of executive functioning and known-group validity such that lower scores differentiated controls from multiple diagnostic syndromes and showed a stepwise decline with increasing disease severity. These results were replicated in a subsample of ALLFTD participants. Taken together, these data suggest the single and pragmatic TabCAT-EXAMINER score is a robust and valid measure of executive functioning across a range of neurodegenerative syndromes.

Several steps were taken to maximize the relevance of the TabCAT-EXAMINER score to a broad research community interested in studying executive functioning. To improve the generalizability of the factor score for use across a range of clinical conditions, the development of the score was conducted using a diagnostically heterogeneous sample (i.e., disease severity, syndrome). Indeed, our results underscore the sensitivity of the TabCAT-EXAMINER score to executive deficits across a wide range of clinical syndromes. Additionally, the factor score was computed using IRT methodology to enhance its use by a variety of researchers with different goals. An important value of IRT is that valid and reliable scores on the same scale can be created using IRT regardless of which tests were administered, enabling researchers to tailor their battery to include only subsets that are pertinent to their study. The TabCAT-EXAMINER demonstrated this feature of IRT through several supplemental analyses that included reduced factor scores generated with fewer components. First, the correlation of the reduced factor score and the full TabCAT-EXAMINER was high, regardless of the number of included items. Variance around that association fell considerably as more items were included. Second, we highlighted individual components within the reduced factor scores. These analyses noted some variability in associations with both the full TabCAT-EXAMINER and the CD CDR®+NACC-FTLD Sum of Boxes across components, but only when the reduced factor included very few components (i.e., two or three of nine). Finally, the TabCAT-EXAMINER demonstrated strong test-retest reliability across time. Taken together, these analyses support a broad use of the TabCAT-EXAMINER, even in studies that do not complete all components.

We also conducted a series of analyses to validate the utility of these tests for FTD participants due to the importance of executive functioning within these syndromes. We first replicated prior investigations with the laptop-based NIH-EXAMINER, finding that the composite score captured executive functioning deficits of FTD, was sensitive to early stages of the disease, and was inversely related to disease severity as indexed by the inverse correlation with CDR®+NACC-FTLD, a measure developed specifically to assess clinical functioning in FTD. In a series of longitudinal analyses, faster rates of decline were observed in those with more advanced clinical severity and participants that showed clinical progression over 6 to 24 months exhibited greater declines in TabCAT-EXAMINER score than those who remained clinically stable. Together, these results support this score as a tool for monitoring disease progression. Across both developmental and validation cohorts, asymptomatic f-FTLD mutation carriers performed worse on average than non-carrier family controls. Although these differences were non-significant and had small effect sizes, that is somewhat expected due to the small number of participants and the heterogeneity of this sample, which comprised three different pathogenic mutations that can present with phenotypic variability. Further study in larger samples is warranted to understand the potential of this score for identifying the earliest cognitive deficits in f-FTLD, which may vary by underlying pathogenic risk. The findings from this study of the TabCAT-EXAMINER score adds further support that digital cognitive tasks are reliable and may serve as effective outcomes for clinical trials (Inan et al., 2020; Staffaroni, Bajorek, et al., 2020; Staffaroni et al., 2024; Staffaroni, Tsoy, et al., 2020).

Lastly, the code to generate these factor scores on new datasets was written in an open-source software, R, and code will be made available at the time of publication to allow interested users to calculate factor scores on their own data. Details about accessing TabCAT software can be found online at: https://tabcathealth.com/. Images of example TabCAT tasks are presented in Supplemental Figure 8.

Our study has some limitations. Although the sample was diagnostically diverse, all participants completed testing in English, and participants were generally highly educated and predominantly white and non-Hispanic. Future studies with non-English speaking and culturally diverse samples are warranted to determine the generalizability of the current findings to other populations. Toward this end, an ongoing study is examining the TabCAT-EXAMINER score within a Latin American Cohort (ReDLat: Multi-Partner Consortium to Expand Dementia Research in Latin America) (Ibanez et al., 2021). Our validation sample primarily comprised FTD participants; additional validation in other neurodegenerative and neurological populations would further demonstrate utility of the TabCAT-EXAMINER.

## Conclusion

Scores on the TabCAT-EXAMINER, a tablet-based battery of executive functioning tests, can be distilled into a single reliable metric with evidence of construct validity and utility for disease monitoring. Since executive functioning is impacted by a range of neurological and medical conditions, this score has broad relevance. These results add to an emerging body of literature supporting the use of digital health technologies for collecting clinical outcome assessments in AD and related dementia research. Future work is needed to understand the relevance of these tests to clinical care and validate their use in different languages and in underrepresented groups.

## Data Availability

The code to generate these factor scores on new datasets was written in an open-source software, R, and code will be made available at the time of publication to allow interested users to calculate factor scores on their own data.

