## Supplement for "Development and validation of the TabCAT-EXAMINER: tablet-based executive functioning factor score for research and clinical trials"

**Neuropsychological measures**

**Convergent Validity**: The following measures are tasks of executive functioning that were not included in the TabCAT-EXAMINER factor.

Digit Span Backwards: The examiner reads a list of digits (e.g., 1-2-3). The participant then repeats those digits, in the backwards order (e.g., 3-2-1). The number of digits increases over time. The outcome measure is the total number of digits a participant is able to repeat in the correct order. This is primarily a task of working memory^1,3^.

Trail Making Test A: Participants connect circles based on the number within each circle. They do so in an increasing order (e.g., 1 to 2 to 3). The outcome measure is the total time it takes to complete the task. In analyses, participant’s scores were reversed to be consistent with other tasks, meaning higher Trails A scores indicate better performance. This task is primarily a task of processing speed^1,3^.

Trail Making Test B: Similar to Trails A, participants connect circles in an increasing order. However, here the circles have numbers or letters. They are asked to switch, in increasing order, between numbers and letters (e.g., 1-A-2-B). The outcome measure is the total time it takes to complete the task. In analyses, participant’s scores were reversed to be consistent with other tasks, meaning higher Trails B scores indicate better performance. This task is primarily a task of inhibition^1,3^.

**Divergent Validity**: The following measures were compared to the TabCAT-EXAMINER with the expectation that associations would not be strongly positive.

Benson Figure Copy: Participants are presented with a simplified version of the Rey-Osterrieth figure that was developed by Frank Benson, M.D. They are asked to copy the figure and their performance is based on a 17-point scale assessing accuracy and placement of design elements. No limit was placed on response time^1^.

Benson Figure Recall: Participants are asked to freely recall the stimulus they copied during the Benson Figure Copy trial. The time delay is approximately 10-15 minutes. Their score ranges from 0 to 17^2^.

Craft Story Recall: Participants are read a short story and asked to immediately recall that story as close to the original words as possible. Then, approximately 20 minute later, they are asked to freely recall this story again. The delayed recall score based on a verbatim scoring method was used in these analyses^3,4^.

Multilingual Naming Test: This task was designed to detect naming impairments in older adults, particularly those diagnosed with Alzheimer’s disease or mild cognitive impairment. Participants are asked to name pictures. The score ranges from 0 to 32, with higher scores indicating better performance^5^.

California Verbal Learning Test-II; Short Form: The California Verbal Learning Test is a 9-item verbal, list-learning task. Participants are read a list of words four times and asked to immediately recall the words after each presentation. Then, after a 10-minute delay, participants were asked to freely recall the word-list. The total number of words correctly recalled after 10-minutes was used in these analyses^6^.

TabCAT Favorites: The participant is shown pairs of verbal and visual stimuli. After the first presentation, they are asked to immediately recall the pairs. They are presented with the same stimuli again, then asked to immediately recall the pairs. Next ,there is a 10-minute delay before they are asked to freely recall pairs without an additional presentation of the stimuli. For the present analyses, the score was the total correct across all three trials (two immediate, one delay).

TabCAT Line Orientation: Participants are shown three lines and ask to identify which two lines are parallel. The difficulty of the task increases based on correct answers and decreases based on incorrect answers by modifying how similar the angle is between the distractor line and the correct response. If the participant answers correctly, the angle difference decreases; if they answer incorrectly the angle difference increases. There are also a series of practice trials used to determine validity (i.e., <80% correct suggests possible invalidity). The outcome score is the average difference in line angle between the distractor and the target line.

**Model Building**: All analyses were completed in R^7^. The first step involved exploratory factor analysis as a means to investigate the overall structure of the data and test the hypothesis that a one-factor model would fit the data well. A scree plot (Supplemental Figure 1) was created to inform the selection of up to nine factors. The scree plot suggested that a single factor model fit the data well, as demonstrated by the sharp decline in the plot: eigenvalue_1-factor_: 5.53; eigenvalue_2-factor_: 1.0; : eigenvalue_3-factor_: 0.71: eigenvalue_4-factor_: 0.61 ; : eigenvalue_5-factor_: 0.34; : eigenvalue_6-factor_: 0.29 ; : eigenvalue_7-factor_: 0.18; : eigenvalue_8-factor_: 0.15; : eigenvalue_9-factor_: 0.13.

The lavaan package^8^ was used to fit a 1-factor confirmatory factor (CFA) mode; it demonstrated poor fit: CFI: .73, TLI: .643, AIC: 10637.7, BIC: 10704, RMSEA: .225. This 1-factor model was compared to a 3-factor model that included a latent factor for the semantic fluency tasks and a latent factor for the lexical fluency tasks. These fluency factors were uncorrelated with the overall executive functioning factor. The 3-factor model notably improved model fit: CFI: .97, TLI: .97, AIC: 10223.3, BIC: 10301, RMSEA: 0.028. A direct comparison between models demonstrated that the 3-factor model offered significantly improved fit (∆χ2 -267.15; p<.001). The structure of this CFA was carried forward into the IRT model which was fitted using the MIRT^9^ package (type: graded; estimation method: Monte Carlo EM). Factor scores for each participant was calculated within MIRT using the “fscores” function (estimation method: expected a-posteriori; quasi-Monte Carlo integration).

**Supplemental Tables and Figures**

| **Supplemental table 1**: Developmental Sample description by genetic status | | | | | | |
| --- | --- | --- | --- | --- | --- | --- |
|  | Full sample | *C9* | *GRN* | *MAPT* | None | Unknown |
| Demographics | | | | | | |
| **Sample Size** | 490 | 41 | 15 | 23 | 132 | 279 |
| **Age** | 56.05 14.98) | 48.73 (12.89) | 57.93 (13.23) | 45.00 (12.53) | 57.12 (13.68) | 57.43 (15.51) |
| **Education** | 16.12 (4.64) | 15.93 (2.52) | 16.00 (3.40) | 19.13 (17.57) | 16.13 (2.94) | 15.90 (2.63) |
| **CDR®+NACC-**  **FTLD** | 3.02 (4.12) | 0.98 (2.56) | 1.63 (3.47) | 2.55 (4.56) | 2.86 (4.24) | 3.52 (4.16) |
| **Male (%)** | 243 (49.7) | 21 (51.2) | 11 (73.3) | 7 (30.4) | 59 (44.7) | 145 (52.2) |
| TabCAT-EXAMINER | | | | | | |
| **Factor Score** | 0.05 (1.03) | 0.63 (0.76) | 0.39 (0.73) | 0.31 (1.11) | 0.30 (0.95) | -0.14 (1.05) |
| **Flanker** | 7.41 (1.43) | 8.10 (0.95) | 7.97 (0.79) | 7.66 (1.58) | 7.63 (1.35) | 7.24 (1.47) |
| **Set Shifting** | 7.00 (1.54) | 7.61 (1.28) | 7.22 (0.89) | 7.49 (1.56) | 7.46 (1.33) | 6.64 (1.60) |
| **Match** | 43.44 (14.47) | 51.48 (8.49) | 46.50 (10.26) | 46.64 (15.34) | 46.74 (12.65) | 40.97 (15.26) |
| **Dot Counting** | 15.34 (6.20) | 17.56 (5.14) | 15.38 (4.50) | 16.25 (7.67) | 15.76 (5.97) | 15.02 (6.36) |
| **Running Dots** | 33.10 (5.16) | 34.34 (5.05) | 31.75 (4.45) | 35.06 (5.51) | 33.52 (5.06) | 32.73 (5.23) |
| **Animals** | 18.75 (8.89) | 23.25 (7.08) | 21.93 (7.54) | 20.36 (9.22) | 20.23 (8.46) | 17.57 (9.06) |
| **Vegetables** | 11.92 (6.26) | 14.07 (5.05) | 13.57 (5.15) | 12.95 (6.15) | 12.62 (6.18) | 11.36 (6.28) |
| **Words (F)** | 12.39 (6.42) | 15.20 (5.02) | 15.07 (5.89) | 13.09 (6.73) | 13.77 (6.24) | 11.28 (6.36) |
| **Words (L)** | 11.45 (5.78) | 13.50 (4.50) | 14.13 (5.96) | 11.33 (5.95) | 12.52 (5.68) | 10.60 (5.73) |
| Diagnosis | | | | | | |
| **CU** | 205 (43.9) | 25 (61.0) | 10 (66.7) | 12 (63.2) | 64 (50.0) | 94 (35.6) |
| **MCI** | 42 ( 6.6) | 6 (14.7) | 2 (13.4) | 2 (10.5) | 5 ( 3.9) | 14 (5.3) |
| **AD** | 2 ( 0.4) | -- | -- | -- | -- | 2 ( 0.8) |
| **bvFTD** | 113 (24.2) | 5 (12.2) | 1 ( 6.7) | 5 (26.3) | 29 (22.7) | 73 (27.7) |
| **CBS** | 10 ( 2.1) | -- | -- | -- | 3 ( 2.3) | 7 ( 2.7) |
| **lvPPA** | 4 ( 0.9) | -- | -- | -- | -- | 4 ( 1.5) |
| **nvPPA** | 25 ( 5.4) | -- | 1 ( 6.7) | 0 ( 0.0) | 9 ( 7.0) | 15 ( 5.7) |
| **PSP** | 9 ( 1.9) | -- | -- | -- | -- | 9 ( 3.4) |
| **svPPA** | 30 ( 6.4) | 1 ( 2.4) | -- | 0 ( 0.0) | 7 ( 5.5) | 22 ( 8.3) |
| **Other** | 38 (11.1) | 4 (9.8) | 1 ( 6.7) | 0 ( 0.0) | 11 ( 8.6) | 22 (8.3) |
| Abbreviations: C9-Chromosome 9 Open Reading Frame 78; GRN-progranulin; MAPT- Microtubule-associated protein tau; None-Negative for genes associated with Familial frontotemporal lobe degeneration; Unknown-Genetic testing not completed, invalid, or missing.  CU-cognitively unimpaired; MCI-mild cognitive impairment; lvPPA- logogenic primary progressive aphasia; AD-Alzheimer’s disease; CBS-corticobasal syndrome; PSP-progressive supranuclear palsy; nvPPA-nonfluent-variant primary progressive aphasia; svPPA-semantic-variant primary progressive aphasia; bvFTD-behavioral-variant frontotemporal dementia; Other-includes those with non-neurodegenerative diseases (e.g., mood) and neurodegenerative diseases with small sample sizes (e.g., Amyotrophic Lateral Sclerosis). | | | | | | |

| **Supplemental table 2:** Validation Sample description by genetic status. | | | | | | |
| --- | --- | --- | --- | --- | --- | --- |
|  | Full sample | *C9* | *GRN* | *MAPT* | None | Unknown |
| Demographics | | | | | | |
| **Sample Size** | 343 | 43 | 12 | 24 | 180 | 77 |
| **Age** | 54.98 (14.59) | 50.95 (13.11) | 51.42 (16.57) | 44.92 (12.14) | 59.16 (14.55) | 55.43 (13.85) |
| **Education** | 16.00 (2.46) | 15.51 (2.12) | 16.33 (2.35) | 15.54 (2.84) | 16.10 (2.64) | 16.25 (2.26) |
| **CDR®+NACC-**  **FTLD** | 1.99 (3.32) | 1.19 (2.37) | 1.00 (3.02) | 1.52 (2.90) | 2.62 (3.83) | 1.91 (3.13) |
| **Male (%)** | 110 (42.8) | 19 (44.2) | 3 (25.0) | 10 (41.7) | 49 (48.5) | 29 (37.7) |
| TabCAT-EXAMINER | | | | | | |
| **Factor Score** | 0.21 (0.91) | 0.50 (0.71) | 0.52 (0.73) | 0.82 (0.69) | 0.08 (1.07) | 0.23 (0.95) |
| **Flanker** | 7.47 (1.32) | 7.73 (1.10) | 7.48 (1.32) | 8.17 (0.98) | 7.43 (1.43) | 7.40 (1.28) |
| **Set Shifting** | 7.31 (1.48) | 7.73 (1.28) | 7.05 (1.01) | 8.39 (1.10) | 7.24 (1.62) | 7.27 (1.46) |
| **Match** | 45.76(12.13) | 48.47 (9.16) | 50.73 (10.08) | 53.96 (6.86) | 42.67 (13.70) | 47.96 (12.32) |
| **Dot Counting** | 16.10 (5.85) | 16.35 (5.74) | 18.10 (6.52) | 18.83 (5.91) | 15.56 (6.47) | 15.70 (4.96) |
| **Running Dots** | 32.86 (5.23) | 32.23 (5.18) | 34.60 (3.75) | 35.60 (4.91) | 32.74 (5.15) | 32.97 (5.69) |
| **Animals** | 21.05 (8.27) | 22.19 (6.67) | 22.75 (7.52) | 23.75 (7.09) | 19.89 (9.10) | 20.82 (8.26) |
| **Vegetables** | 13.15 (6.09) | 14.33 (5.03) | 14.33 (3.14) | 13.58 (4.99) | 12.65 (6.84) | 12.82 (6.25) |
| **Words (F)** | 13.46 (6.12) | 15.09 (4.62) | 16.33 (7.19) | 14.88 (5.58) | 13.13 (6.64) | 12.10 (5.82) |
| **Words (L)** | 12.41 (6.16) | 13.98 (4.18) | 13.92 (7.35) | 14.17 (6.58) | 11.64 (6.25) | 11.75 (6.49) |
| Diagnosis | | | | | | |
| **CU** | 140 (54.5) | 27 (62.8) | 10 (83.3) | 15 (62.5) | 46 (45.5) | 42 (54.5) |
| **MCI** | 21 ( 8.2) | 4 ( 9.3) | 1 ( 8.3) | 5 (20.8) | 6 ( 5.9) | 5 ( 6.5) |
| **AD** | 2 ( 0.8) | 0 ( 0.0) | 0 ( 0.0) | 1 ( 4.2) | 0 ( 0.0) | 1 ( 1.3) |
| **bvFTD** | 40 (15.6) | 8 (18.6) | 1 ( 8.3) | 3 (12.5) | 18 (17.8) | 10 (13.0) |
| **CBS** | 4 ( 1.6) | 0 ( 0.0) | 0 ( 0.0) | 0 ( 0.0) | 2 ( 2.0) | 2 ( 2.6) |
| **lvPPA** | 1 ( 0.4) | -- | -- | -- | -- | 1 ( 1.3) |
| **nvPPA** | 14 ( 5.4) | -- | -- | 0 ( 0.0) | 9 ( 8.9) | 5 ( 6.5) |
| **PSP** | 8 ( 3.1) | -- | -- | 0 ( 0.0) | 5 ( 5.0) | 3 ( 3.9) |
| **svPPA** | 14 ( 5.4) | -- | -- | 0 ( 0.0) | 7 ( 6.9) | 7 ( 9.1) |
| **Other** | 13 ( 5.1) | 4 ( 9.3) | -- | 0 ( 0.0) | 8 ( 7.9) | 1 ( 1.3) |
| Acronyms: C9-Chromosome 9 Open Reading Frame 78; GRN-progranulin; MAPT- Microtubule-associated protein tau; None-Negative for genes associated with Familial frontotemporal lobe degeneration; Unknown-Genetic testing not completed, invalid, or missing.  CU-cognitively unimpaired; MCI-mild cognitive impairment; lvPPA- logogenic primary progressive aphasia; AD-Alzheimer’s disease; CBS-corticobasal syndrome; PSP-progressive supranuclear palsy; nvPPA-nonfluent-variant primary progressive aphasia; svPPA-semantic-variant primary progressive aphasia; bvFTD-behavioral-variant frontotemporal dementia; Other-includes those with non-neurodegenerative diseases (e.g., mood) and neurodegenerative diseases with small sample sizes (e.g., Amyotrophic Lateral Sclerosis). | | | | | | |

| **Supplemental table 3:** Sample description and TabCAT-EXAMINER scores by CDR®+NACC-FTLD Global Score subgroups within the validation cohort | | | | | | | |
| --- | --- | --- | --- | --- | --- | --- | --- |
|  | **Full sample** | **CDR®+NACC-FTLD Global Score** | | | | | |
|  |  | **0** | **0.5** | **1** | **2** | **3** | **Comparison** |
| Demographics | | | | | | | |
| **Sample Size** | 343 | 146 | 49 | 43 | 18 | 1 |  |
| **Age** | 54.98 (14.59) | 47.86 (13.35) | 62.49 (11.83) | 66.02 (8.25) | 65.67 (9.54) | 59.00 |  |
| **Education** | 16.00 (2.46) | 15.89 (2.33) | 16.14 (2.94) | 16.42 (2.30) | 15.56 (2.43) | 16.00 |  |
| **Male (%)** | 110 (42.8) | 48 (32.9) | 26 (53.1) | 25  (58.1) | 10 (55.6) | 1 |  |
| TabCAT-EXAMINER | | | | | | | |
| **Factor Score** | 0.21 (0.91) | 0.81 (0.51) | 0.20 (0.77) | -0.69  (0.81) | -1.32 (0.71) | -1.79 | 0>0.5>1>2 |
| **Flanker** | 7.47 (1.32) | 8.02 (0.93) | 7.32 (1.29) | 6.53  (1.39) | 5.80 (1.52) | -- | 0>0.5>1 |
| **Set Shifting** | 7.31 (1.48) | 7.97 (0.99) | 7.44 (1.38) | 5.90  (1.78) | 5.70 (0.98) | -- | 0>0.5>1 |
| **Match** | 45.76 (12.13) | 53.20 (6.87) | 44.33 (10.04) | 34.36 (9.93) | 26.31 (15.05) | 30.00 | 0>0.5>1 |
| **Dot Counting** | 16.10 (5.85) | 18.27 (4.74) | 15.17 (5.20) | 13.10 (6.40) | 7.77 (4.87) | 2.00 | 0>0.5>1 |
| **Running Dots** | 32.86 (5.23) | 34.16 (4.67) | 33.00 (4.56) | 30.14 (6.00) | 23.43 (5.13) | -- | 0>0.5>1>2 |
| **Animals** | 21.05 (8.27) | 24.82 (5.50) | 21.59 (7.23) | 13.42 (7.20) | 8.22 (5.54) | 2.00 | 0>0.5>1>2 |
| **Vegetables** | 13.15 (6.09) | 16.03 (4.21) | 12.59 (5.40) | 7.69  (5.51) | 4.78 (4.45) | 0.00 | 0>0.5>1>2 |
| **Words (F)** | 13.46 (6.12) | 16.30 (4.53) | 12.94 (4.70) | 8.42  (5.85) | 4.56 (3.99) | 2.00 | 0>0.5>1>2 |
| **Words (L)** | 12.41 (6.16) | 15.10 (5.05) | 11.57 (4.94) | 7.77  (5.68) | 4.56 (3.75) | 1.00 | 0>0.5>1>2 |
| The “Comparison” column indicates significance of comparisons (T-test or Chi^2^ test) between CDR®+NACC-FTLD Global score groups. Statistical comparisons involving the CDR®+NACC-FTLD Global Score = 3 group were not computed due to small sample size. Means, (standard deviations), and, when applicable, sample size [n], are presented. Male provides raw count (percent). | | | | | | | |

**Supplemental Figure 1:** Skree plot

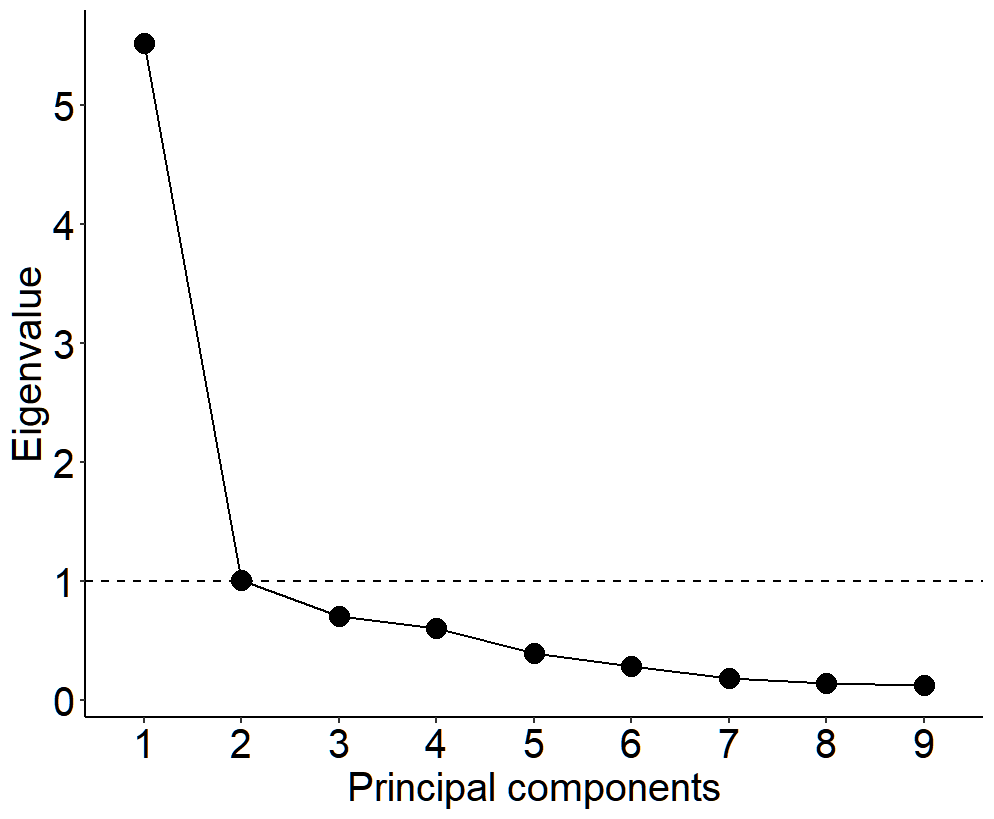

Displays the number of principal components (X-axis) and eigenvalues (Y-axis). A horizontal dashed line has been placed at the “elbow” of the scree plot to indicate the location of a sharp decline in eigenvalues.

**Supplemental Figure 2:** TabCAT-EXAMINER distribution for the full development sample and by CDR®+NACC-FTLD Global Scores

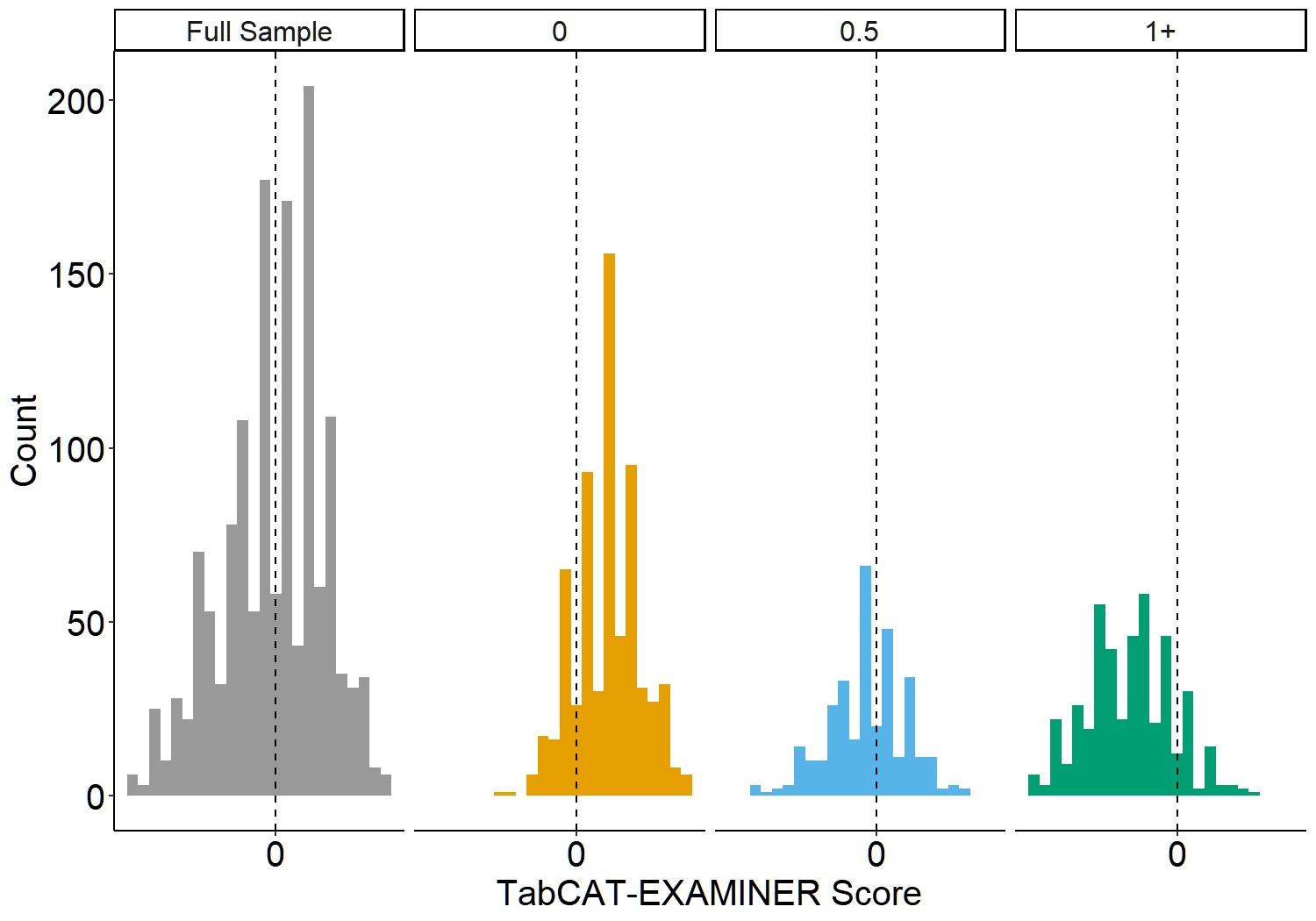

Each histogram details the number of participants on the y-axis and the TABCAT-EXAMINER score on the x-axis. Histograms are presented for the full sample (gray) and those with NACC®+NACC-FTLD Global Score of 0, 0.5, and 1+.

**Supplemental Figure 3**: Correlations between the TabCAT-EXAMINER score created with all available tests and the composite score created with select numbers of subtests

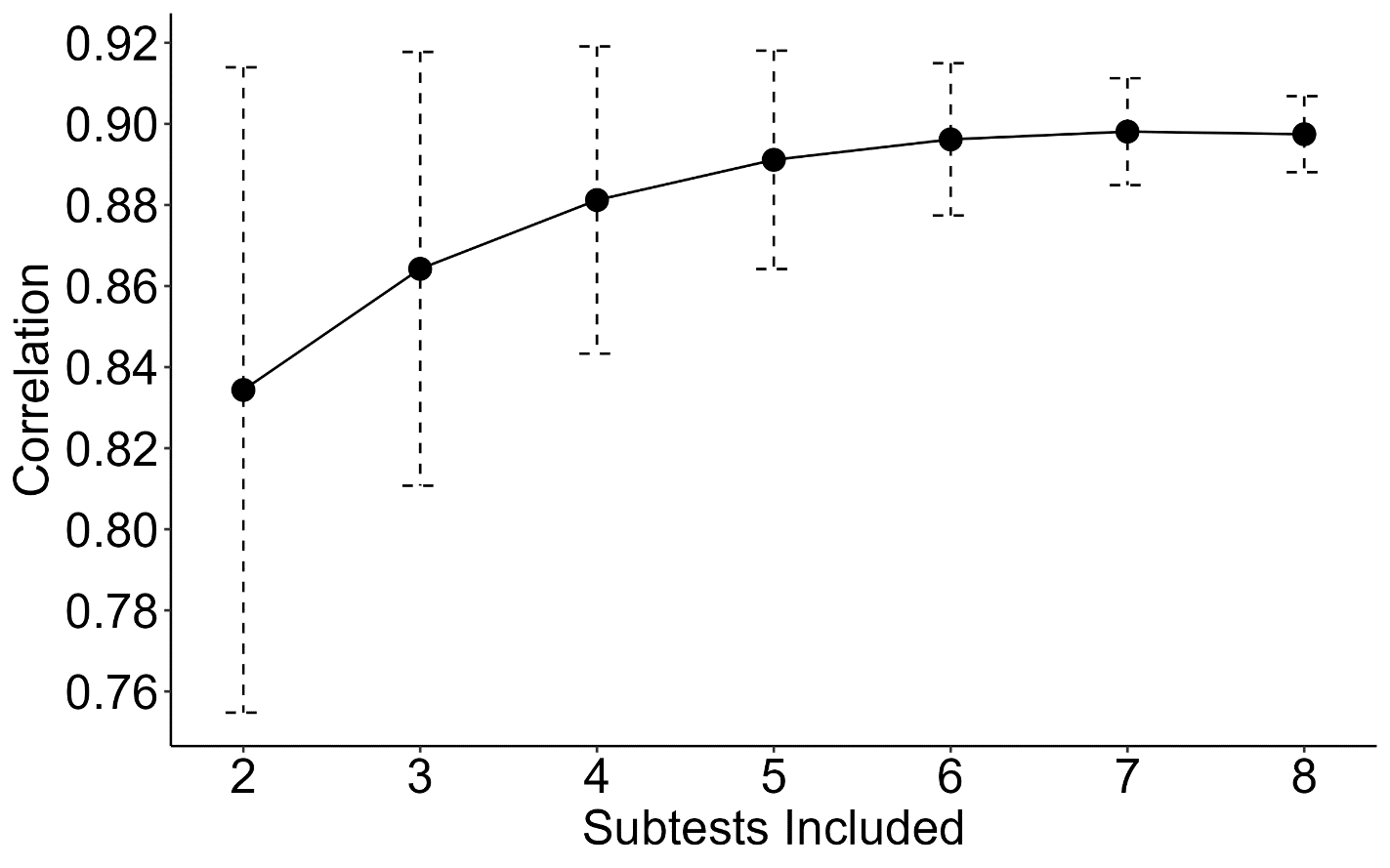

The X-axis indicates the number of subtests included in the creation of the factor score, ranging from two of nine to eight of nine. A total of 502 reduced factor scores were created, semi-randomly drawing, without replacement, from all potential combinations of components. The Y-axis presents the average correlation (with confidence interval) for a component created with that number of subtest and the full, nine-subtest TabCAT-EXAMINER score.

| **Supplemental Figure 4:** Correlations between reduced factors scores and the full TabCAT-EXAMINER by specific component and number of items in model | |
| --- | --- |
| 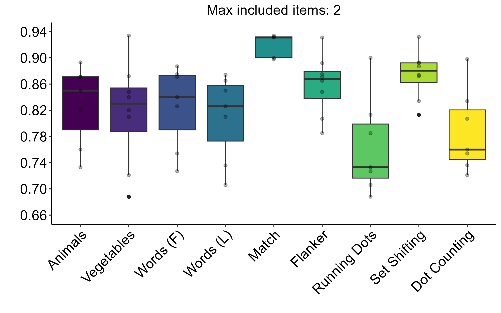 | 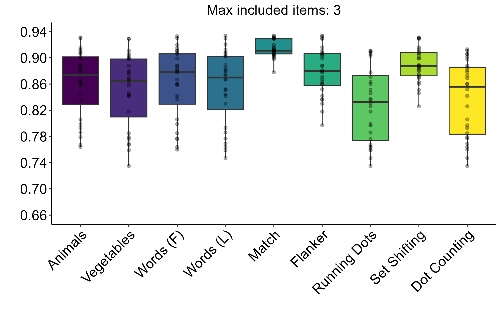 |
| 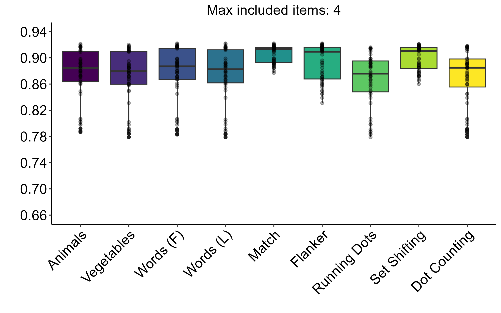 | 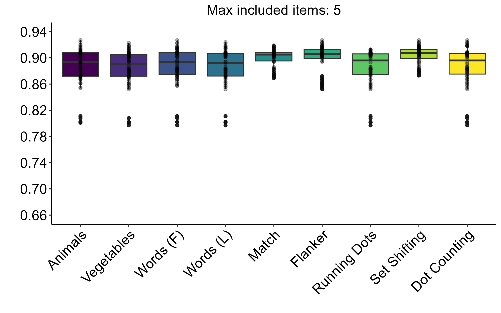 |
| 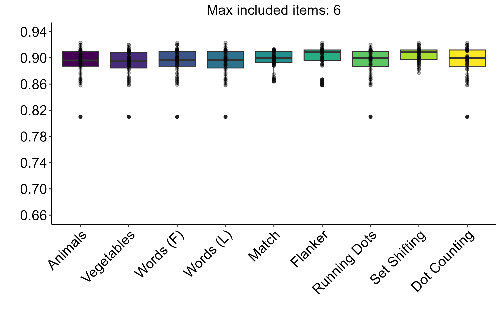 | 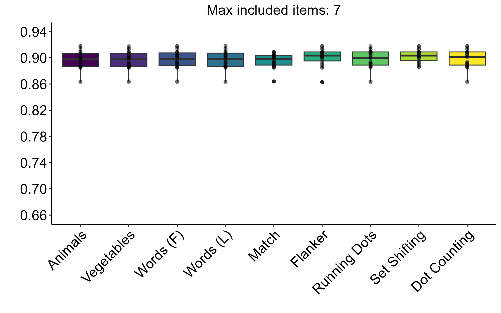 |
| Seven figures present the association between reduced factor scores and the full TabCAT-EXAMINER within models defined by the number of components included (e.g., two of nine, three of nine etc.). The Y-axis within each figure presents the Pearson correlations between each reduced factor score and the full TabCAT-EXAMINER. The X-axis indicates which component was required in the model. For example, the “Animals” column within the first figure (“max included items: 2”) indicates models with two components, where one component must be the Animals variable. The boxplot indicates the average association and 95 percent confidence interval. The dots indicate the correlation for each included model. Models with eight items were completed but the figure was excluded for brevity as results were similar to models with seven items. | |

| **Supplemental Figure 5:** Correlations between reduced factors scores and the CDR+NACC-FTLD Sum of Boxes by specific component and number of items in model | |
| --- | --- |
| 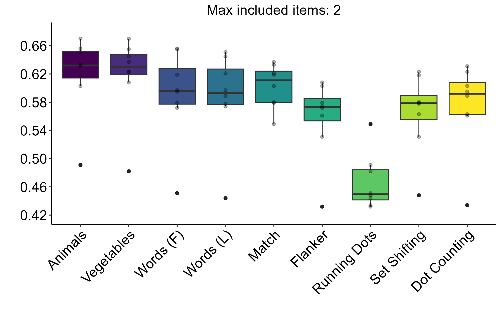 | 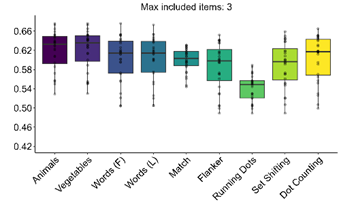 |
| 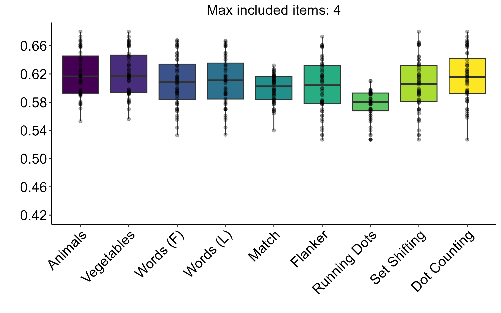 | 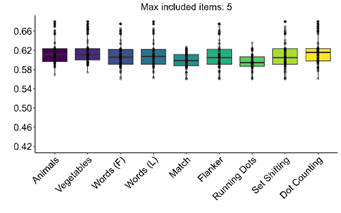 |
| 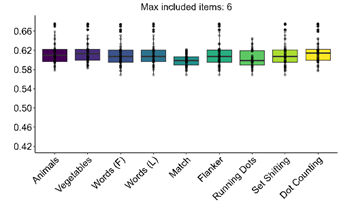 | 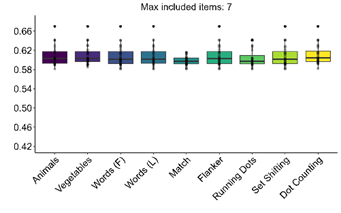 |
| Seven figures present the association between reduced factor scores and the CDR+NACC-FTLD Sum of Boxes within models defined by the number of components included (e.g., two of nine, three of nine etc.). The Y-axis within each figure presents the Pearson correlations between each reduced factor score and the CDR+NACC-FTLD Sum of Boxes . The X-axis indicates which component was required in the model. For example, the “Animals” column within the first figure (“max included items: 2”) indicates models with two components, where one component must be the Animals variable. The boxplot indicates the average association and 95 percent confidence interval. The dots indicate the correlation for each included model. Models with eight items were completed but the figure was excluded for brevity as results were similar to models with seven items. | |

**Supplemental Figure 6:** Baseline TabCAT-EXAMINER performance by clinical syndrome within the validation sample

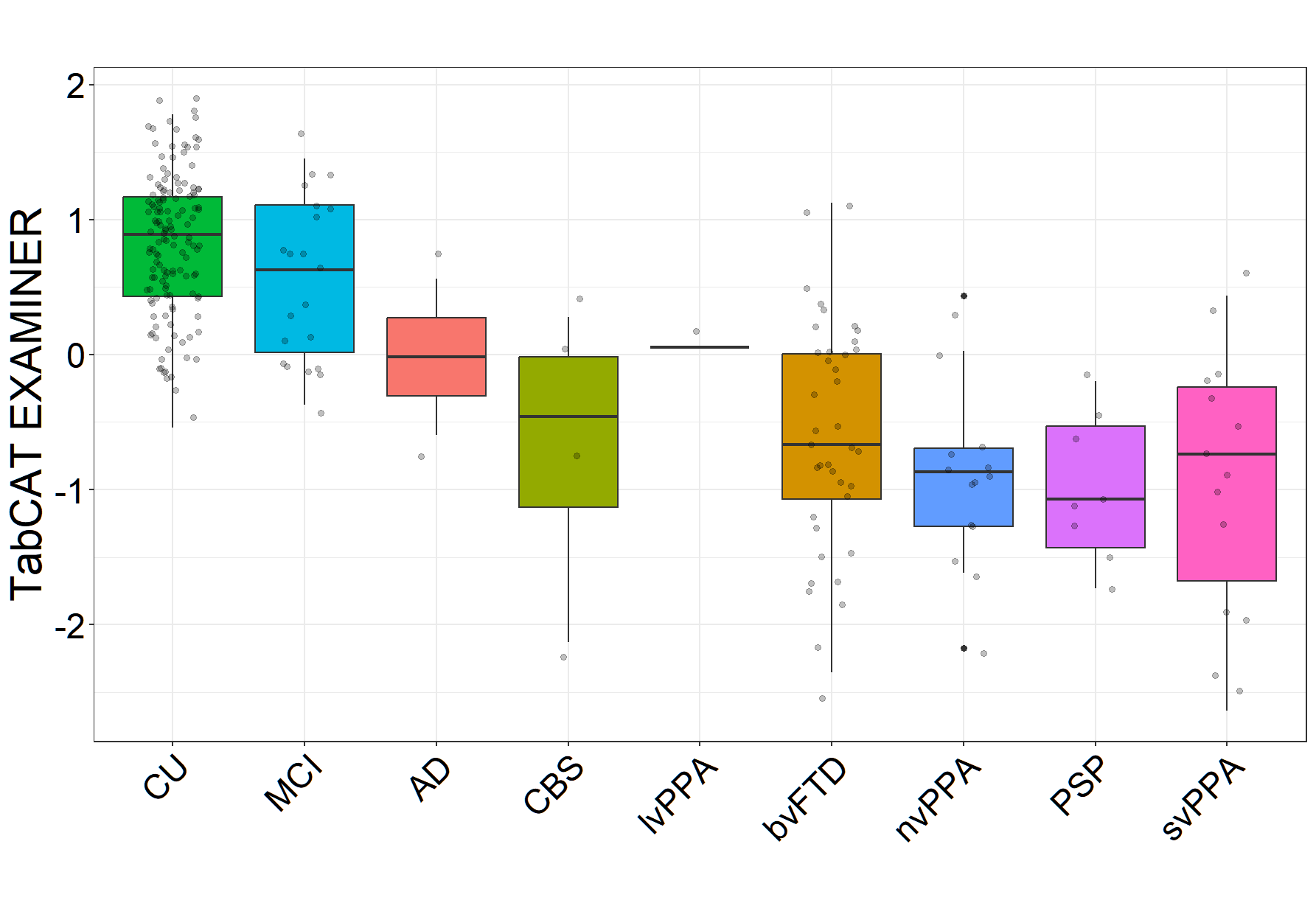

Boxplots present TabCAT-EXAMINER scores for diagnostic syndrome groups.

Abbreviation: CU-cognitively unimpaired; MCI-mild cognitive impairment; lvPPA- logopenic primary progressive aphasia; AD-Alzheimer’s disease dementia; CBS-corticobasal syndrome; PSP-progressive supranuclear palsy; nvPPA-nonfluent-variant primary progressive aphasia; svPPA-semantic-variant primary progressive aphasia; bvFTD-behavioral-variant frontotemporal dementia

**Supplemental Figure 7**: Test-Retest Reliability within cognitively unimpaired participants in the validation sample

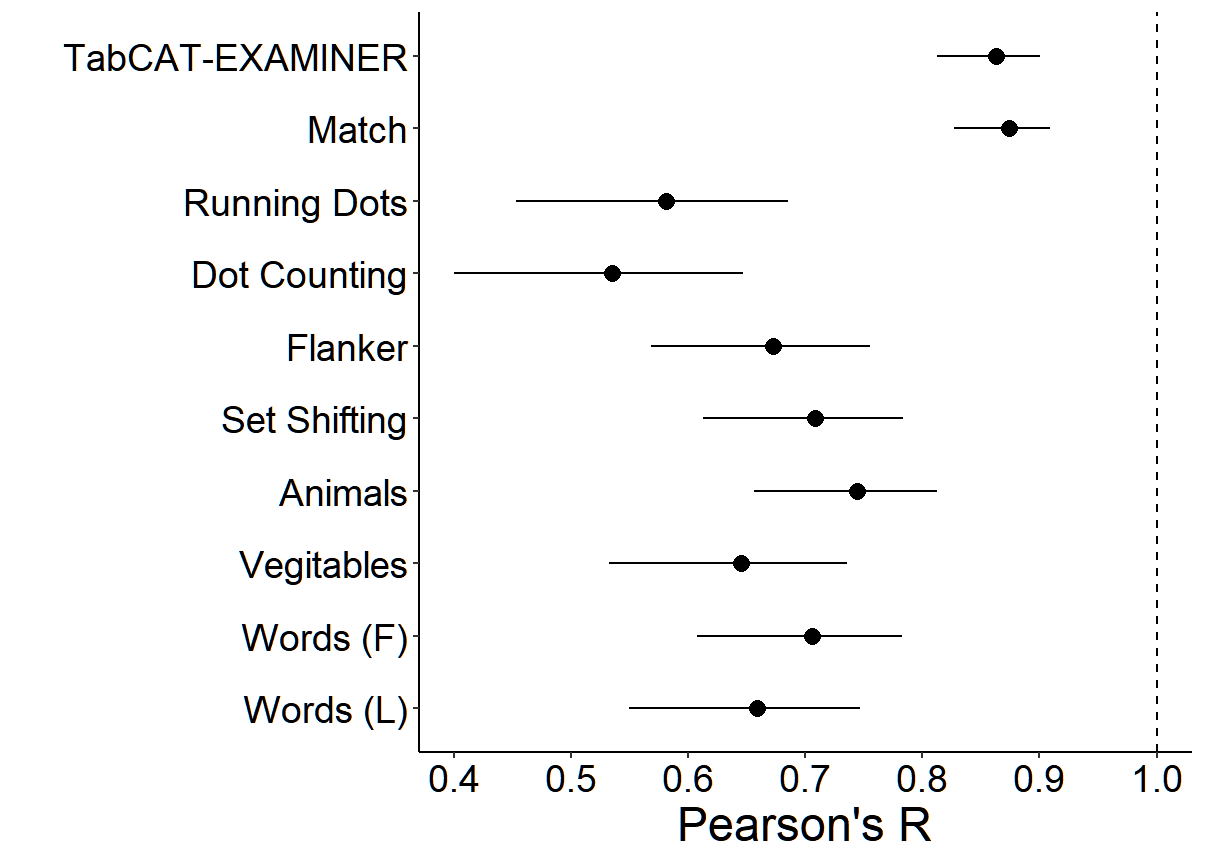

Presents Pearson’s correlations and 95 percent confidence intervals between baseline and follow-up TabCAT-EXAMINER scores as well as component measures. The sample included 146 participants who were cognitively unimpaired (CDR®+NACC-FTLD Global score = 0) at both time points.

| **Supplemental table 8:** TabCAT-EXAMINER tablet-based measures | | | |
| --- | --- | --- | --- |
| **Dot Counting**  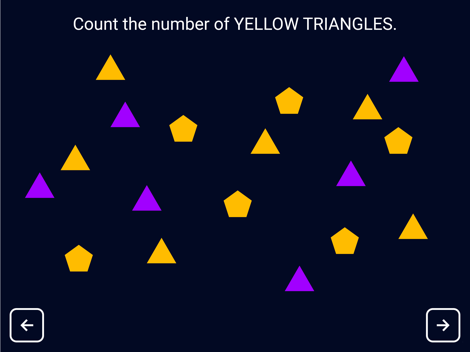 | **Running Dots**  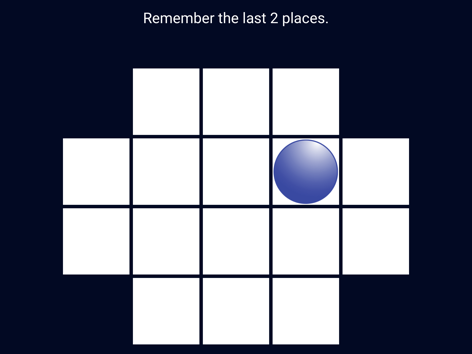 | | **Set-Shifting**  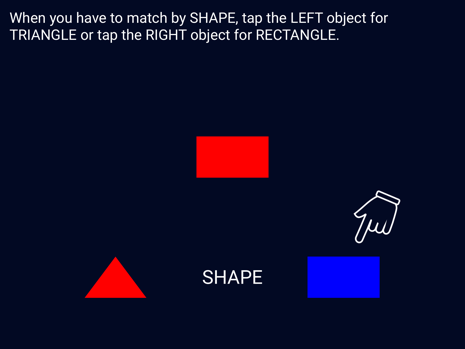 |
| **Match**  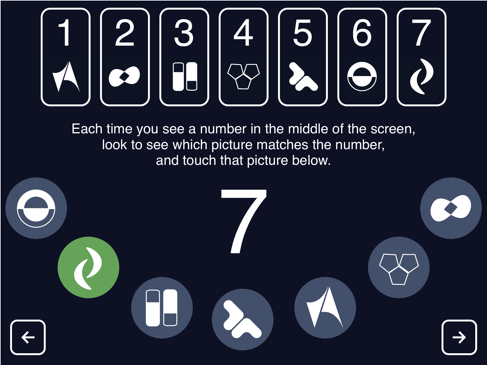 | | **Flanker**  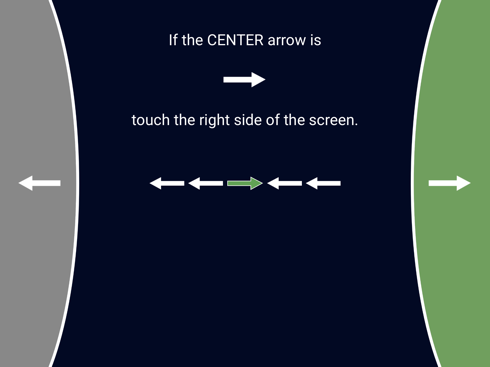 | |
| Provides sample images for each of the TabCAT measures used in the TabCAT-EXAMINER factor. Due to test security concerns, images above have been altered and do not present exact stimuli. More information on the TabCAT tasks can be found at: <https://tabcathealth.com/>. The fluency tasks are part of the Unified Data Set (UDS)^1^ and the original NIH-EXAMINER^2^. | | | |
